# Comparison of SARS-CoV-2 Exit Strategies Building Blocks

**DOI:** 10.1101/2020.04.23.20072850

**Authors:** Elad Barkan, Smadar Shilo, Yeela Talmor-Barkan

**Affiliations:** Weizmann Institute of Science, Rehovot Israel; Rabin Medical Center (Beilinson and Hasharon Hospitals), Petach Tikva, Israel

## Abstract

We consider and compare various exit strategy building blocks and key measures to mitigate the current SARS-CoV-2 pandemic, some already proposed as well as improvements we suggest. Our comparison is based on a computerized simulation integrating accumulated SARS-CoV-2 epidemiological knowledge. Our results stress the importance of immediate on-symptom isolation of suspected cases and household members, and the beneficial effects of prompt testing capacity. Our findings expose significant epidemic-suppression differences among strategies with seemingly similar economic cost stressing the importance of not just the portion of population and business that is released, but also the pattern. The most effective building blocks are the ones that integrate several base strategies - they allow to release large portions of the population while still achieving diminishing viral spread. However, it may come with a price on somewhat more complex schemes. For example, our simulations indicate that a personal isolation of 4 days once every two weeks, for example a long weekend (Fri-Mon) self-isolation once every two weeks, while protecting the 5% most sensitive population would reduce R well below 1 even if ten percent of the population do not follow it. This kind of integrated strategy can be either voluntary or mandatory and enforced. We further simulate the contrasting approach of a stratified population release in a hope to achieve herd immunity, which for the time being seems inferior to other suggested building blocks. Knowing the tradeoff between building blocks could help optimize exit strategies to be more effective and suitable for a particular area or country, while maximizing human life as well as economic value. Given our results, we believe that pandemic can be controlled within a reasonable amount of time and at a reasonable socio-economic burden.

## Introduction

Coronavirus disease 2019 (COVID-19) caused by severe acute respiratory syndrome coronavirus 2 (SARS-CoV-2) was declared as a pandemic by the World Health Organization ^1^. Unfortunately, no vaccine is available at this time. Therefore, mitigation actions are being taken to minimize both the adverse effects of the pandemic on the economy and the spread of the virus. Currently taken measures are mostly social distancing, isolation of cases and suspected cases, contact tracing, higher levels of hygiene, facial masks wearing, and mass population quarantines which cause vast socio-economical damage ^2–6^. Going forward there’s a need to easen some of the measures to allow economic activity, while minimizing viral health damage.

One extreme approach is a stratified release of population, lower risk first. The idea is that lower risk individuals would contract the virus, recover and gain immunity, thus slowing the epidemic rate for future uninfected individuals. The hope is that by the time higher risk individuals are released, the infection probability is greatly reduced and thus infection of the higher risk group is largely reduced. Hence the term “*Herd Immunity*”. Care is taken to release at a rate that would not overload the healthcare system.

In the other extreme there are mitigation building blocks based on “social distancing”. These are deliberate measures taken to restrict, slow and limit the spread of the virus such that only a very small number of individuals will end up infected until the disease is eradicated or a vaccination is available. Social distancing measures could reduce the probability of contracting the virus given a contact, for example using facial masks, or reducing the rate of contacts of infected and susceptible individuals - thus reducing the spread of the virus ^4–7^. These measures typically include keeping a large portion of the population at home either constantly or intermittently. The challenge is how to reduce the economic and social cost of such measures to an acceptable level while controlling for viral spread.

The mid-term goal is keeping R_t_<1 through social distancing measures, where R_t_ indicates at every time (t) how many secondary infections are caused on average by a single infected case. R_t_>1 means the disease is spreading at an exponential growth rate as each case causes more than a new single case, and R_t_<1 means the disease is diminishing as each case causes less than a new single case. The special case of R_t_=1 or very close to one is not so theoretic, and suggests the disease continues at a fixed rate of new daily cases. In such a case, a short intervention can reduce the number of daily cases and then we can continue with the same measures, but in a lower rate of new daily cases. Reduction of new daily cases could also assist in contact tracing, if done manually - which further slows the epidemic.

R_t_ at time zero is called the basic reproduction number R_0_. For SARS-CoV-2 the exact number is under active research and it could change from one country to another or given different environmental conditions ^8–10^, with many estimated values in the range of 2.2-3.2 which means each case causes 2.2-3.2 new cases, unless people have already been infected and cannot get infected twice. When social distancing measures are taken, the reproduction rate changes, and it is then denoted by R instead of R_0_.

Social distancing measures have a potentially high price tag attached to them, both economically and socially. Therefore, once the rate of new infections is low enough, we want to keep R_t_<1 but close to 1 with measures that cost the least. The situation is constantly changing and keeping R_t_<1 but close to 1 is a moving target - for example there are constant changes in human behaviour, public attention and public resilience, weather ^11,12^ changes, local outbreaks and so on and all could affect R_t_. We need to dynamically try different measures so we keep R_t_<1 but close to 1 while probing and estimating the current situation. Therefore, we could occasionally cross the line to R_t_>1, which means growing infection rates until we take actions to reduce R_t_. The price paid for social distancing interventions is also in the difficulty of implementing them. Complex measures could mean less compliance and less success in implementation and eventual effectiveness.

As the current goal is to keep R_t_ around 1 and the situation is constantly changing, information and response speed are the key. The consequences of quick feedback are priceless compared with alternatives. If the feedback loop for an action taken is 14-21 days, and during this time the virus could be back propagating at doubling the number of cases every 3-4 days. We could be expecting 3-7 sequential multiplications, i.e., 8-128 fold increase (2^3^ to 2^7^) in the number of new infections, and following a similar increase in ICU beds. Therefore, lack of prompt feedback, typically through mass-testing leads to a need to maintain a large spare capacity. Maintaining spare capacity has a large cost associated with it, both monetary and possibly in human life. We propose how to promptly estimate the number of new infections and thus gain a short response time to actions taken.

In the first part of the paper, key mitigation measures are suggested. Our findings stress the importance of self-isolation upon symptoms until a SARS-CoV-2 test result, and further isolation of household members if no fast testing is available or if a test returns positive for the virus.

The second part of the paper focuses on comparison between building blocks in terms of their effectiveness in controlling R_t_<1, and effectiveness in terms of percent of population business-day that is released. Surprisingly, there is a large difference in epidemic-control effectiveness of strategies, while having similar economic costs by releasing the population to a similar extent. The key is not just the portion of the population that is released, but also the release pattern. This finding suggests that exit strategies could be optimized for both viral spread and economic value. The more successful strategies were the more complex ones, integrating several ideas. Gaining understanding of the tradeoff is important as it could assist in building an exit strategy suited for a specific area, in a way that is cost effective and communicable to the public so compliance would be high. For each building block, we give the resulting reduction in R as indicated by our simulation.

For example, to reduce R well below 1 it suffices that every person enters a four-day isolation at their pre-chosen time within each two weeks period while protecting the 5% most sensitive population. This suggestion works even if people would take their isolation over a long weekend once every two weeks (Fri-Mon), and even if ten percent of the population do not follow it. It works even if in addition every person in a household chooses a different isolation period, and no isolation is kept among household members.

The reason such a strategy works is three-fold:

1. Interaction is greatly reduced 4 out of 14 days - 28.5% of time is significant in terms of virus spread.
2. Infectiousness onset 3 days on average after contracting the virus. Those that contract the virus during their 10 release days have a good chance to enter their pre-planned isolation shortly after or before they become infectious and thus infect less.
3. As symptoms typically onset on day 5 from contraction, those that contract the virus during their 10 release days have good chances that symptoms onset would occur during their pre-planned isolation or shortly after. With self isolation upon symptoms they are likely to be removed from further infecting others.

Together with general social distancing measures such as self-isolation on symptoms and wearing facial masks while outside of home, the result is a reduction of R well below 1.

We give special consideration to the herd immunity approach. Given the knowledge that has already accumulated, our results indicate that for now social distancing is expected to be a significantly more effective approach than herd immunity, with significantly less casualties across all age groups.

## Part I: Key Mitigation Measures

A word of caution. Our results are based on a SEIR agent-based simulator, which we built based on Israeli population structure of nine million people, and based on existing knowledge on SARS-CoV-2 epidemiological behavior. The simulations are performed in a 1-day iteration cycle simulating a period of one year. We distinguish between infection within a household and outside, as existing literature shows the virus spreads significantly in familial infection clusters ^13^. Various exit-strategy building blocks are fed into the simulator so their outcome can be assessed given the existing knowledge. The simulation is executed 10 times over each set of parameters, but with different random choices. Standard deviation of the results in these runs are given in time-based figures, when a point value is given it corresponds to the mean result. The exact structure and full list of assumptions is given in the supplementary at the end of this paper.

### Proposal 1: Immediate household self-isolation upon first symptom

We propose that the single most cost-effective epidemic control is <u>immediate</u> self-isolation of the <u>entire</u> household upon <u>the first symptom</u> ^14^ suspected as related to coronavirus of someone in the household. Release from isolation is only after a negative laboratory result of the first person in the household to have shown a symptom, and provided that the other members of the household do not show any symptoms related to coronavirus.

In our simulations, after this measure is taken, R dropped from R_0_=3.03 to R=1.54. This is a dramatic drop by a factor of 1.54/3.03 = 0.508. This drop is compared to the second-best epidemic control measure: immediate self-isolation on the first symptom of a person (without other household members), which results in R=2.2, a 2.2/3.03=0.726 factor drop. If the symptomatic person self-isolates on symptoms but we wait with household isolation until the virus test for the symptomatic person returns positive, and assuming a 5-day delay we get a much inferior situation with R=1.65. The effect size in this case between an immediate household isolation to a delayed one 1.54/1.65 = 0.93 is comparable to the effect of facial masks with protection factor of 10% 2.83/3.03=0.93, see Figure 2. These few days post symptoms and before diagnosis could be critical for reducing further infections directly or through other household members ^15^. See Figure 1. If vast and prompt tests are in place, the difference between household isolation and personal isolation upon symptoms diminishes, as household members are assumed to enter isolation upon a positive test for one household member, and get checked themselves.

**Figure 1:**
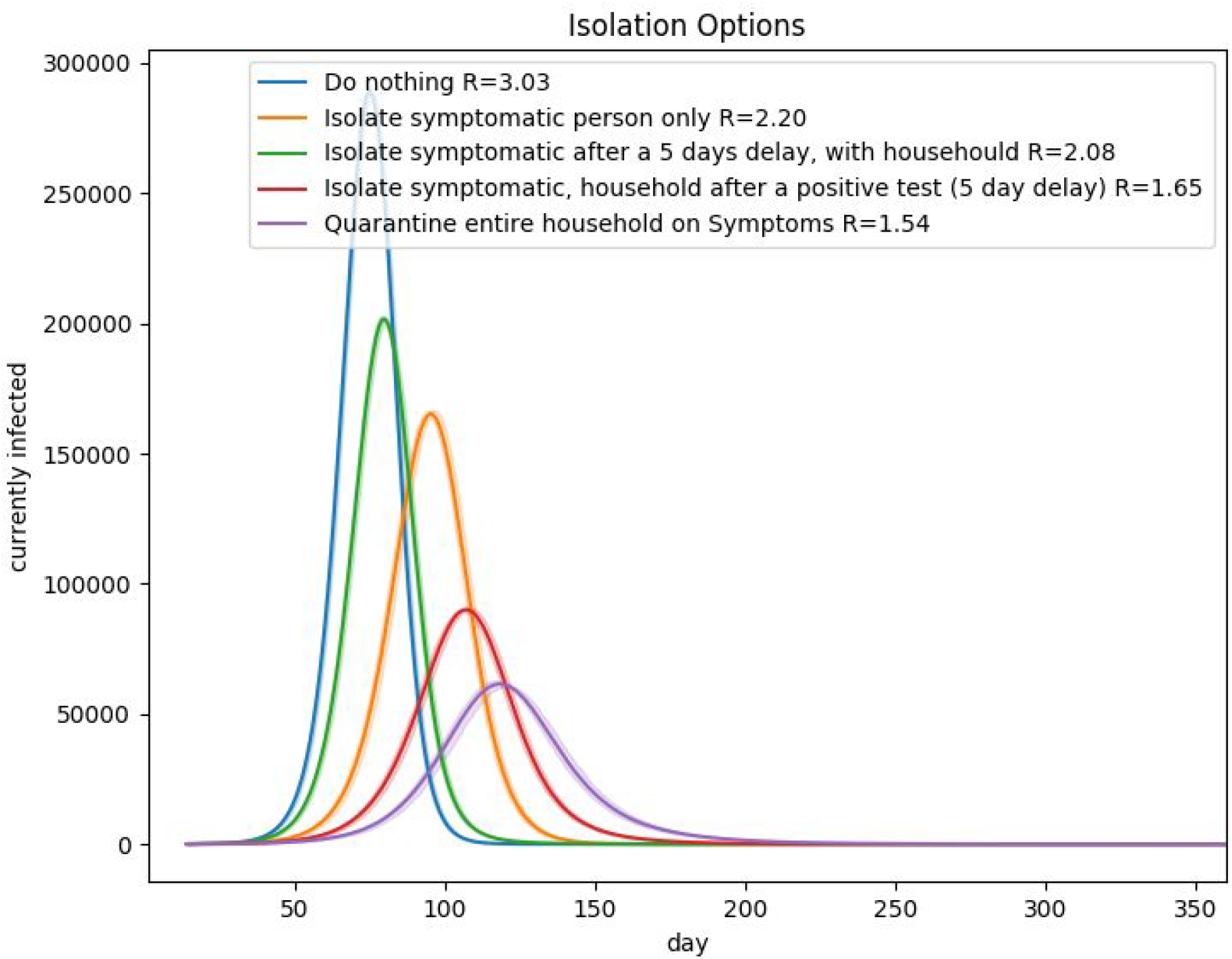
Comparison of isolation options. Simulation is based on Israeli population N=9,096,440. Five options: no isolation, self-isolation of symptomatic person and household only after a a test returns positive 5 days later, self-isolation of a person upon the first symptom, self isolation of a person upon the first symptom - household members join after a test returns positive 5 days later, self-isolation of the entire household upon the first symptom.

**Figure 2:**
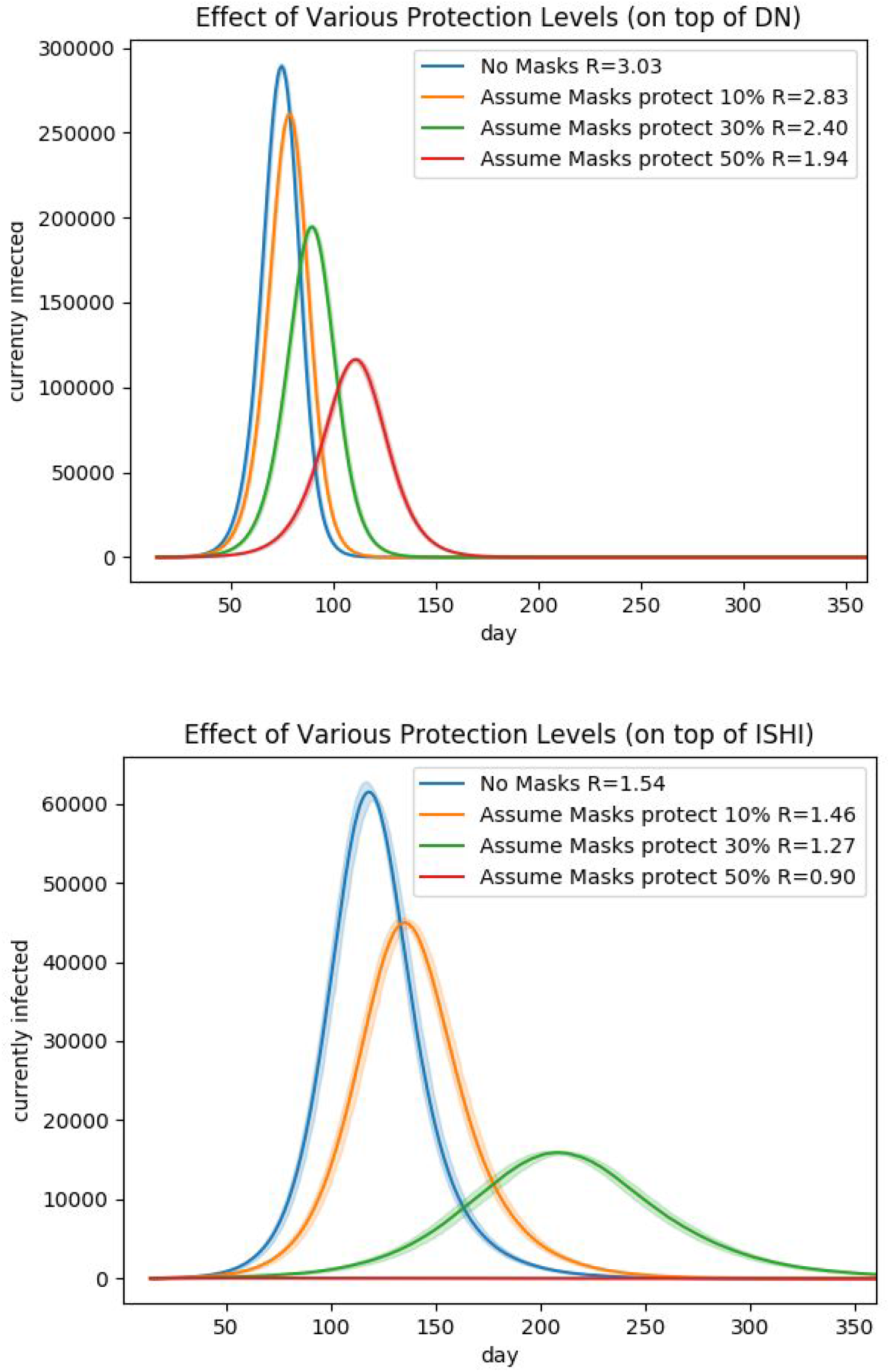
“What if” Comparison of various protection levels (masks, hygiene, etc.). At this point in time, the effect of such measures on SARS-CoV-2 transmission is unknown. **Top**:no isolation. **Bottom**: isolation of households on first symptom

Immediate self-isolation of the entire household achieves an immediate stop to further spreading of the virus of both possibly affected but asymptomatic members and possibly other affected but presymptomatic household members. Without fast and prompt testing it would probably not be enough to isolate just the symptomatic person, as research based on experience in China shows that a large proportion of the infections occur within households ^13,16^, see particular example ^17^. Patients are pre-symptomatic in the first few days after infection while still being possibly infecting. Furthermore, it’s estimated that a vast portion of the infections are made by pre-symptomatic or asymptomatic ^18–23^. Social distancing measures taken outside the household, reduces non-household infections. The declining infection rate between households means that relatively, more infection occurs within a household. It is also interesting to notice that areas with larger households are likely to suffer from a higher R even if all other parameters are the same. Our simulations show that a self household isolation, even on the basis of fever alone of one of the members, has a dramatic effect in preventing the spread of the epidemic, and should be used together with any other measures taken ^24^.

Self-isolation upon symptoms dramatically shortens the spreading period. A study on Chinese experience in Shenzhen shows it took an average of 4.6 days after symptoms onset to isolate an infected person, which was reduced to 2.7 days if the person was isolated due to contact tracing and not symptoms ^13^.

One implementation option is that every person of a household shall self-test for fever every day, with possible daily online compulsory reporting of body temperature as well as onset of other symptoms. Body temperature could also be checked in key public locations or entry points. We estimate the daily number of new people with fever by assuming 0.3 percent of the adult population shows fever signs of above 38 Celsius at any given day ^25^, and dividing by an average conservative duration of a non-COVID-19 fever disease of 1-4 days^26–28^. Therefore, an order of 7,500 to 30,000 isolations shall occur daily and thus this number of daily tests for this purpose would be needed for a country the size of Israel about 9 million people. Only if the first symptomatic household member is found to be infected, a test would be carried out to the other family members - so the extra tests due to infected household members is expected to be insignificant compared to the background symptomatic non-SARS-CoV-2 cases.

An improvement to reduce the amount of tests would be using machine learning algorithms utilizing additional external data to release a household from self-isolation without the need for a laboratory test for the virus. Such external data can include the prevalence of coronavirus in the surrounding area, prevalence of other infectious diseases in the area, workplace, or by information on personal location voluntarily collected and stored on household members’ mobile phones ^29^ etc.

### Proposal 2: Focus testing capacity on self-isolated households

Focus testing capacity on quickly testing self-isolated households. This approach simultaneously serves three important purposes in parallel:

1. **Quick and accurate measure of actual coronavirus spread** - as on average symptoms onset about 5.2 days after infection ^30^, and households enter immediate self-isolation, immediate testing gives an accurate and near real-time measure of virus spread in the community, and could prevent the need for population survey-testing for the virus.
2. **Effective contact tracing -** Given self-isolated households contain the population with the highest risk of being infected that could have already continued unidentified infection chains, there’s importance in locating those that are really infected and trace their contacts to isolate them. When testing is done quickly enough, we have better chances of isolating the non-household individuals that got infected by the detected cases, before they infect others. We note that we did not include contact tracing effects in our simulations, so the total combined effect of isolation and quick testing could be even stronger than we report.
3. **Public Compliance and fast reporting -** when the logic behind self-household isolation is shared with the public, and the public knows that they would be released in a short time from self-isolation if they didn’t contract coronavirus, the public is more likely to comply with the instructions, and with compliance the virus spread would reduce. As symptoms might be mild to begin with, people might delay the request for a test. In the simulations we assume this delay would be half a day on average as we conservatively assumed people would only comply when having fever symptoms, and fever can be tested when entering the workplace, schools, and in a variety of other public places. If testing is fast enough, it suffices to isolate only the symptomatic person until results arrive.

In the unfortunate scenario that there is not enough testing capacity to cover all the new symptomatic candidates, the importance of the proposed self-household isolation is even greater, as effective contact tracing would not be possible and infection has a significant familial base ^13^. In such a case, sample testing of the isolated households might still provide a good proxy for the spread of the infection.

It should be noted that not testing all the isolated population could lead to increasing public in-acceptance. Perhaps when the number of new infections is high and it’s likely the symptoms are due to SARS-CoV-2 infection, people would accept a delay in testing. Probably less when the number of new infections shall drop to a few a day, and the chances of justified isolation shall be minimal yet crucial in preventing relapse.

### Proposal 3: Vaccinate population for common influenza towards the upcoming winter

Many countries regularly recommend the entire population (unless contraindicated) to vaccinate against seasonal influenza towards the winter ^31^. We suggest strengthening this recommendation towards the winter this year. Seasonal influenza could cause coronavirus-like initial symptoms ^32^, and vaccination shall reduce the background noise of seasonal influenza symptoms, which could reduce the number of needed tests. In addition, reduction in influenza reduces pressure on the healthcare system and keeps the population in a healthier starting point. In short, reduction in influenza allows to focus scarce resources on the coronavirus front. Likewise, the standard vaccination schedule should be kept and its importance communicated to the public.

### Proposal 4: Facial Masks and Hygiene Measures

As the cost of facial masks is low, and absent information, counties are tempted to instruct the population to wear facial masks in addition to increased hygiene ^4–6^. The hope is that these measures could reduce random infection by SARS-CoV-2 ^2,3^. Whether these alone could stop the epidemic and what their contribution would be, if any, is unknown at this time. Nevertheless, in Figure 2, we provide three simulations, assuming facial masks and hygiene measures reduce infection outside home by 10%, 30%, and 50% respectively. Note we assume the measures are taken only while outside home, so household infection remains the same. See further analysis in the supplementary.

## Part 2: Improvement and Comparison of Exit Strategies Building-Blocks

We compare the efficiency of several key exit strategies building blocks, comparing how much they are efficient in stopping the epidemic in terms of R, while in relation to how effective they are the amount of average released business days they allow for the population. We summarize results of the partial release options in Figure 3.

**Figure 3:**
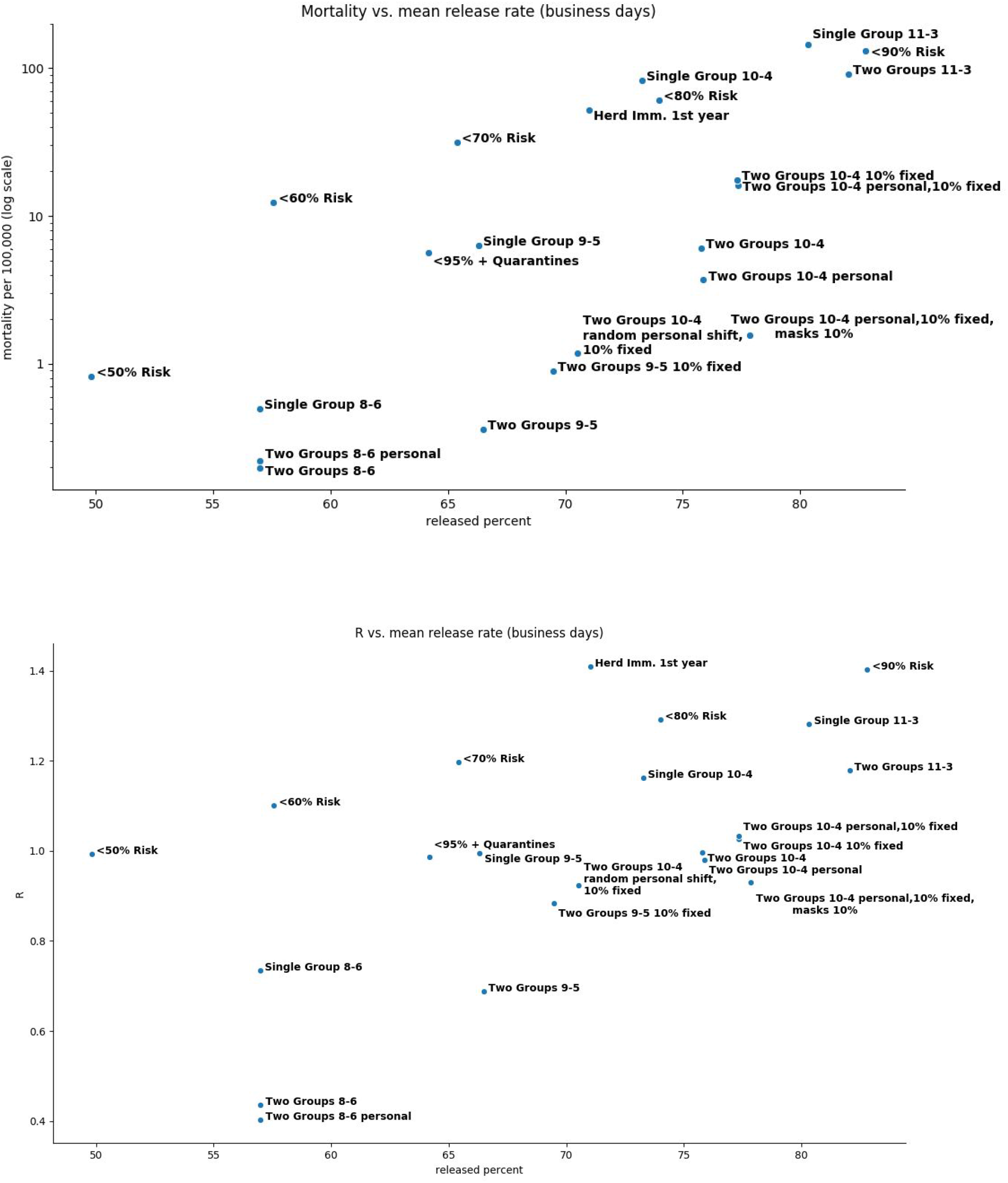
**Top**: mortality per 100,000 vs *released percent*. **Bottom**: R vs. *released percent. released percent* - the average per-person percent of business days (Mon-Fri) in which a person is released based on a building block in a 1-year period.

Exit strategies building blocks we consider:

1. **Do nothing - let the virus swipe the population (DN)**
2. **Immediate self-isolation upon symptoms (ISI)**
3. **Self-isolation of entire households only after a positive test (5 days delay)**
4. **Immediate self-household-isolation upon symptoms (ISHI)**
5. **Immediate self-isolation of symptomatic, household joins after a positive test (5 days delay)**
6. **DN + wearing facial masks when outside home (DN+masks)**
7. **ISHI + wearing facial masks when outside home (ISHI+masks)**
8. **Intermittent Release**. Intermittent release of population, see ^33^
9. **Release certain percent** of the population, quarantine the rest. The released group does not change across days.
10. **Release 95% of the population with 1-week quarantines as needed:** Release 95% healthiest by households, then a week quarantine is entered if virus spread causes above 2e^-5^ (2 in 100,000) true positive new household isolations daily averaged for a week.
11. **Improved Two-Group intermittent release**. Suggested below.
12. **Improved Two-Group intermittent with fixed 10% release**. Suggested below.
13. **Random shift with fixed 10% release**. Suggested below. A special category of its own of deliberate exposure while controlling exposure order:
14. **Herd immunity** - described below.

In all strategies except DN, ISI, and herd immunity, we isolated households upon the first symptom. For all simulations, we assumed leakage of infection from the released population to the quarantined population, such that being quarantined reduces the probability of infection from outside sources by a factor of 100 compared to the released population. While we cannot justify the exact factor, the need to model leakage is undoubted as there’s always some level of incompliance, special needs that require going outside, need to buy supplies, human mistakes etc.

Personal risk prediction was based on a mocked personal coronavirus mortality prediction - see supplementary, and for a household as the average risk for the household members. Alternatively, risk can be evaluated by age based on SARS-CoV-2 estimated infection fatality rate. In all simulations except DN, ISI, ISHI,DN+masks and ISHI+masks, we kept the top 5% highest-risk household in isolation to protect them. In practice, other protection measures for sensitive populations are required, together with sensitive locations - these are outside the scope of the current paper.

### Improved intermittent release

In this family of strategies we greatly improve an earlier proposed family of strategy ^33^. In these strategies, we release the population based on 14-day cycles, for a varying number of consecutive days depending on the particular variant. In the remainder of the days in the cycle, the previously released population is set on quarantine which we refer to as *washout period*. During the washout period, some of those who contracted the virus in the preceding release days would become symptomatic. Hence, they will be “washed out” from further infecting others before the next cycle begins. Another advantage of this family of strategies is that during the washout days R_t_ is smaller due to the reduction in interactions.

We suggest a few improvements that together with ISHI make a big difference:

1. **Dynamic and gradual number of release days**, **vs. quarantine days**. The object is to add as many release days while keeping R_t_ below 1. Based on measured near real-time R_t_ (discussed above), if R_t_ is below 1, and the load on the healthcare system is reasonable and the number of new true-positive daily household isolations is below desired threshold, we add a release day on expense of a quarantine day in a cycle. If R is growing above 1, we deduct a release day on the expense of a quarantine day in each 14-days cycle. We manage to significantly increase the number of release days in a cycle from 4 suggested in the original idea ^33^ to 9, thanks to our ISHI isolation procedure, and despite that we assumed R_0_=3.03. Additional measures could further increase the number of release days while keeping R_t_ < 1.
2. **Two-group approach reduces R and allows more release days**. We suggest to divide the population into two groups A and B, whose quarantine days are disjoint but their release days can be joined. The population can be divided household-wise. The strength of this improvement is that while it keeps the washout periods in place, it further reduces infection on days that only one group is released. On each day that only one group is released, the number of interactions decrease and limit further infection in the group. Thus, R is effectively reduced. Of the possible day-divisions between the groups, we suggest divisions which maximizes the number of business days in an equal manner between the groups. We give several examples in the tables below, and note that with this adaptation, we can improve to 10 workdays out of a 14 days cycle, while keeping R below 1. In other words a long weekend (Fri-Mon) once every two weeks suffices to reduce R below 1 under our model assumptions.
3. **Division to groups on a personal rather than household basis** One of the possible difficulties in implementing a two-group approach is dividing into two groups based on households rather than on a person by person basis. Surprisingly, our simulations show that the difference in the basis of division is not significant in respect to R, while it could make a big difference in the ease of implementation. Therefore, we suggest this improvement of dividing to groups based on a personal basis. This basis can also be set by the workplace, as long as it makes sure the division is such that it lowers in-workplace physical interaction.
4. **Two-Group approach with a fixed release** Although effective, a two-group approach could appear more difficult compared to other building blocks, mainly as some employees are essential and are needed at their workplace more than their group designates them or the group division is violated due to poor compliance. Therefore, we present an option allowing a fixed release rate in parallel to the two groups. For example, a fixed 10% of the population could be released all the time, regardless of the group division. This improvement makes the scheme more real, at the cost of somewhat elevated R - see Figure 4 for 80% business days. As we see, while the cost in terms of R is significant, rising from R=0.98 for Two groups 10-4 days division on personal based group-division, to R=1.03 with additional 10% fixed population release - it’s still very close to 1. Even with just the addition of facial masks (or hygiene) at 10% protection it would bring us to R=0.93. This improvement allows for a long weekend (Fri-Mon) once every two weeks with division on a personal basis, with 10% of the people in complete noncompliance - a dramatic improvement in implementability while still keeping R below 1.
5. **Random shift with a fixed release** The maximum level of freedom can be given, allowing people to choose their 4 isolation days along 14 days, allowing 10% noncompliance and no masks. If the choice is random, it shows better performance in terms of R (R=0.92) compared to the same building block with two groups (R=1.03), and in terms of business days it’s 70.52% vs. 77.31% respectively. Business days average is lower as isolation can fall over business days instead of weekends - but this is a personal choice. For R, the division is sensitive to people coordinating their isolation period to align and maximize interaction. In such a case we shall end up in a situation similar to two groups, or in the worst case to a single group R value. However, as circles in life are diverse, we assume it would be nearly impossible for the public to completely synchronize their isolation days.

**Figure 4:**
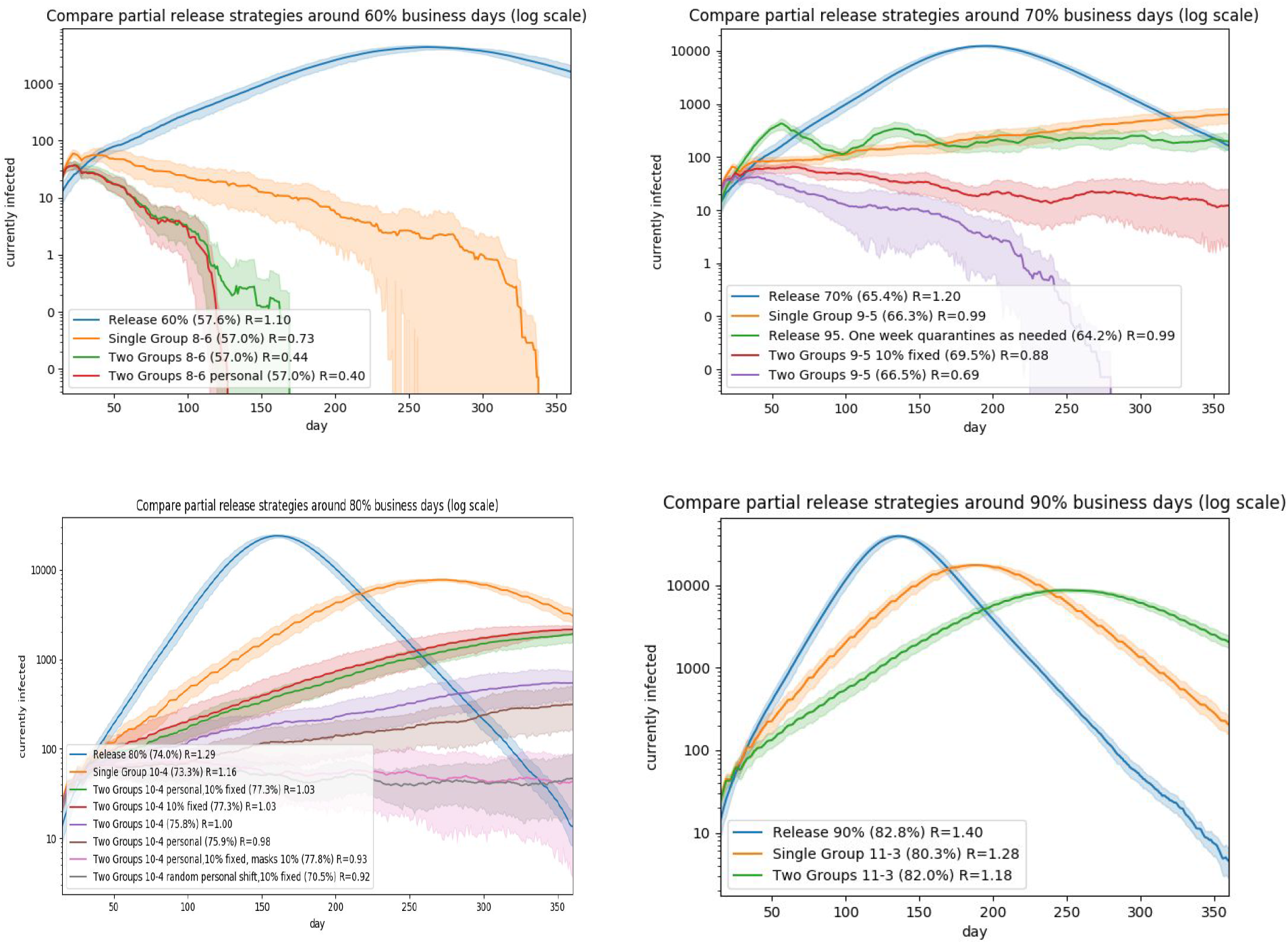
Comparison of partial release strategies based on percent of released population business-days (graphs are in log scale).

This strategy can be either voluntary, or enforced for example by having individuals fill-in online in advance their chosen self-isolation days for future 14-day periods, and this strategy building block can come on top of other strategies such as wearing masks - leading to an even better epidemiological result.

We give a few examples:

**Two Groups 8-6**: 8 release days followed by 6 quarantine days:

| Group | M | Tu | W | Th | F | Sa | Su | M | Tu | W | Th | F | Sa | Su | Avg | Mon-Fri |
| --- | --- | --- | --- | --- | --- | --- | --- | --- | --- | --- | --- | --- | --- | --- | --- | --- |
| A |  | + | + | + | + | + | + | + | + |  |  |  |  |  | 57.1% | 60% |
| B | + | + | + |  |  |  |  |  |  | + | + | + | + | + | 57.1% | 60% |
| Total | $\frac{1}{2}$ | Full | Full | $\frac{1}{2}$ | $\frac{1}{2}$ | $\frac{1}{2}$ | $\frac{1}{2}$ | $\frac{1}{2}$ | $\frac{1}{2}$ | $\frac{1}{2}$ | $\frac{1}{2}$ | $\frac{1}{2}$ | $\frac{1}{2}$ | $\frac{1}{2}$ | 57.1% | 60% |

**Two Group 9-5**: 9 release days followed by 5 quarantine days:

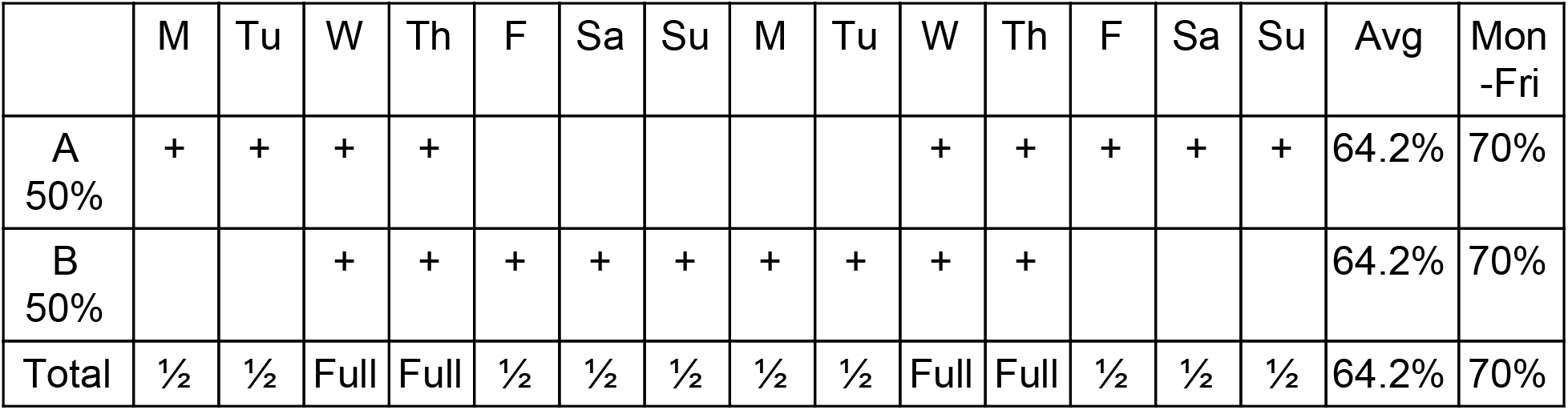

|  | M | Tu | W | Th | F | Sa | Su | M | Tu | W | Th | F | Sa | Su | Avg | Mon-Fri |
| --- | --- | --- | --- | --- | --- | --- | --- | --- | --- | --- | --- | --- | --- | --- | --- | --- |
| A<br>50% | + | + | + | + |  |  |  |  |  | + | + | + | + | + | 64.2% | 70% |
| B<br>50% |  |  | + | + | + | + | + | + | + | + | + |  |  |  | 64.2% | 70% |
| Total | $\frac{1}{2}$ | $\frac{1}{2}$ | Full | Full | $\frac{1}{2}$ | $\frac{1}{2}$ | $\frac{1}{2}$ | $\frac{1}{2}$ | $\frac{1}{2}$ | Full | Full | $\frac{1}{2}$ | $\frac{1}{2}$ | $\frac{1}{2}$ | 64.2% | 70% |

**Two Group 10-4**: 10 release days followed by 4 quarantine days:

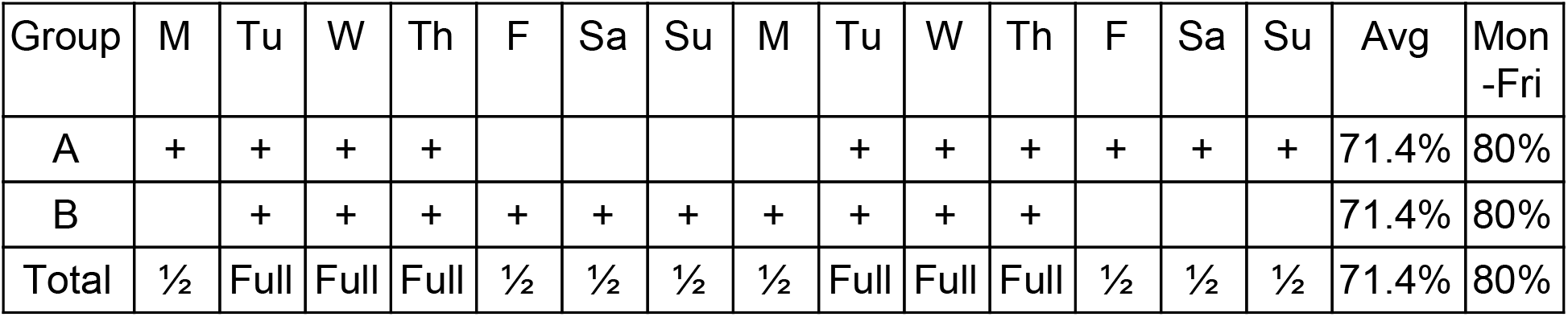

**Two Group 11-3**: 11 release days followed by 3 quarantine days:

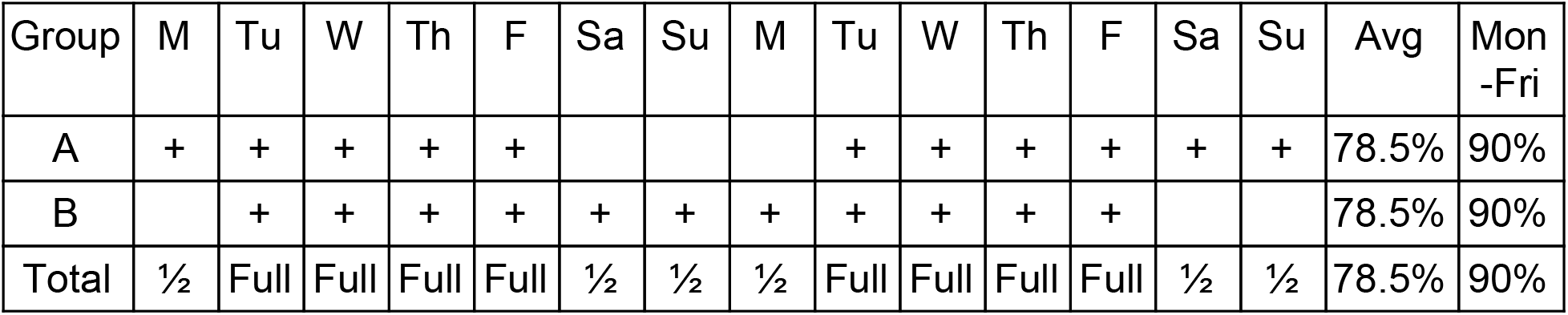

### Protection of population at Risk - Expected Mechanical Ventilation by Household

| Percentile stratas by household risk profile | Percent requiring ventilators, household-risk by fitted predictor (see below) | Percent requiring ventilator, scored by age group of household risk profile based on age-only<sup>24,34</sup> |
| --- | --- | --- |
| 0-25% | 0.045% | 0.059% |
| 25-30% | 0.0811% | 0.092% |
| 30-35% | 0.0972% | 0.1127% |
| 35-40% | 0.1177% | 0.14% |
| 40-45% | 0.146% | 0.166% |
| 45-50% | 0.185% | 0.264% |
| 50-55% | 0.237% | 0.339% |
| 55-60% | 0.30% | 0.5% |
| 60-65% | 0.39% | 0.74% |
| 65-70% | 0.51% | 1.22% |
| 70-75% | 0.712% | 1.65% |
| 75-80% | 1.03% | 2.58% |
| 80-85% | 1.59% | 3.99% |
| 85-90% | 2.74% | 5.0% |
| 90-95% | 5.97% | 10.29% |
| 95-100% | 24.5% | 10.55% |

The higher-risk percentiles take an enormous amount of medical resources relative to their size, where the top 5% risk-group take a 544-fold more resources vs. the lower risk group (using the fitted predictor for risk assessment - see supplementary), or 178-fold more resources if we use age alone for risk assessment. The resources go hand in hand with their risk, which demonstrates why this population needs special protection also from the standpoint of resource usage, especially in the top 5%-10% percentiles.

### Herd Immunity

We discuss herd immunity separately from other strategy building blocks, as it is the only strategy that aims at increasing infection rates - even if only in certain age groups in an attempt to protect the rest and perhaps the economy. All other strategies are focused on preventing further infection. As such, herd immunity has one big advantage over all other strategies - immunity from relapse, under the assumption that a person cannot get infected twice. This advantage is believed to be only a temporary one, as the general belief is that if we wait long enough - there will be a vaccine that will provide immunity without the need to get large portions of the population sick, or that global efforts of social distancing and testing will eventually eradicate the disease.

The logic behind the herd immunity approach is that the lowest risk households are released first, and as they are low-risk they shall have a reduced demand rate of healthcare resources and reduced casualties rate. The low-risk groups shall then be first to contract the virus, recover and gain immunity. Thus, the virus transmission is reduced as those that recover do not infect others and cannot get sick again. Further groups of increasing risk are then released too, in a measured rate as not to exceed healthcare capacity. The extremely high-risk groups shall be released last once the epidemic is over.

Therefore, we release the lowest-risk 25% immediately, and release further 5% of the population every two weeks until ICU is at 1% capacity. As a result, ICU utilization peaks and slightly overshoots, and we continue releasing higher risk groups every two weeks once ICU capacity is back to at least 50% capacity again.

The herd immunity approach tries to infect as many of the released population as fast as possible in the beginning, so we did not isolate symptomatic households at start. We employ the ISHI procedure to slow virus transmission only after 60% of the population has been released.

We depict simulation results for herd immunity in Figure 5 vs. a Two Group 10-4 strategy with division into two groups on personal basis, allowing up to 10% complete noncompliance and using facial masks outside hume assuming 10% protection. The later strategy allows more release business days on average in a 1-year period - 77.8% vs. 71% for herd immunity.

**Figure 5:**
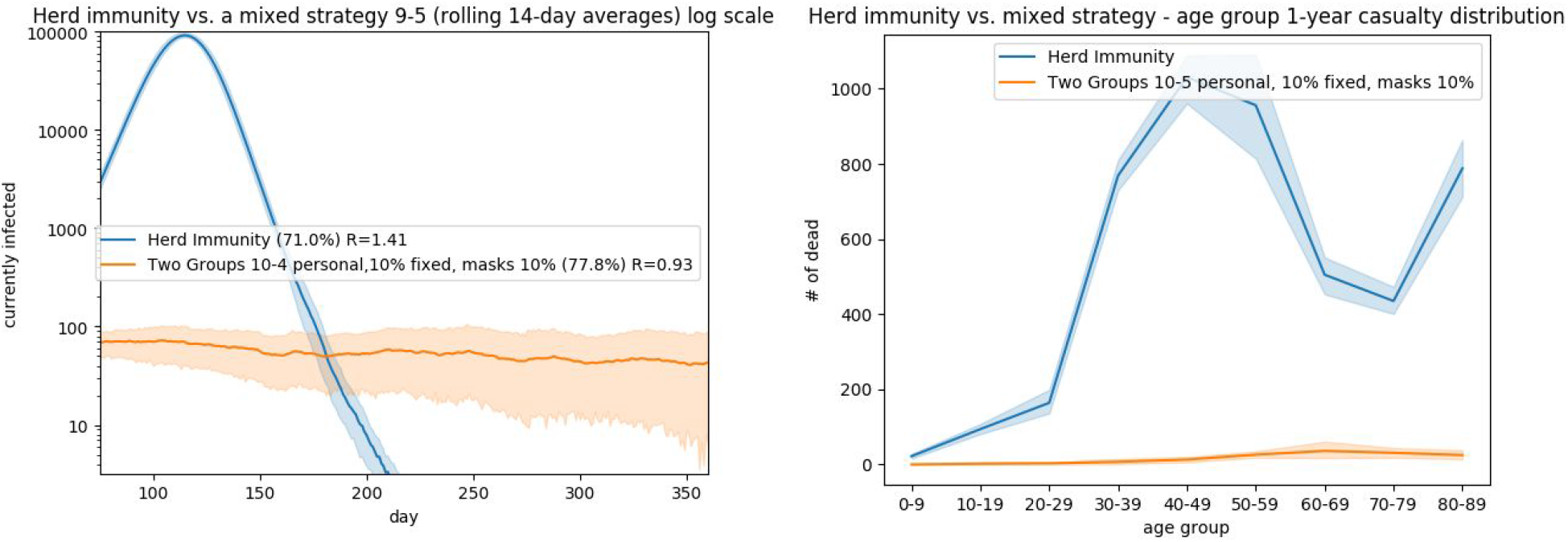
1-Year Comparison of herd immunity strategy vs. Two-Group 10-4 division on personal basis, 10% noncompliance, masks protection at 10%

### Herd immunity should probably wait

There are several points that push away from a herd immunity strategy at this stage:

1. **Too many need to get infected**. Herd immunity is by far an outlier strategy with the largest amount of infections. From what we currently know, we don’t expect herd immunity to appear before over 60% of the population contracts the virus. While it’s reasonable to assume that a person shall not contract the virus twice in a short period, it’s still an unknown risk. Even for a small country the size of Israel of about 9 million people, it means about 5,400,000 million people need to get infected - and just for Israel. Infecting that many people could result in mistakes being multiplied in very large numbers, and almost definite leakage of infection to more sensitive populations that cannot be really protected in 100% of total isolation for months - resulting in many casualties.
2. **Closes the door to potential luck**. We might get lucky and have the summer eliminate the epidemic, as happened with some other viral infections - or other measure being successful. We might have an effective medicine that will allow people to get infected without serious consequences. With herd immunity at this point - we won’t enjoy this luck, or enjoy it in a limited way.
3. **Inefficient, costly for the economy and inhumane, as it will take too long to implement**. We take for example Israel which has a relatively young and healthy population structure. It is expected to take many months of quarantine given the limited capacity of the healthcare system, keeping a large portion of the population under quarantine for a prolonged period of time - which can be considered inhumane. We simulate a year ahead, and give the herd immunity approach the advantage that we ignore death after a 1-year period. As can be seen in Figure 3 and Figure 5, we have other strategies that can, at least potentially, hold the situation with minimal number of infections and casualties for for a relatively low cost, until we have a vaccine for example.
4. **Too many will die and die young**. While mortality rate is becoming clearer and much lower than initial estimates, it’s still relatively high ^34^. Herd immunity approach could lead not only to higher mortality than alternatives, but out of the casualties a high proportion would be young. At this time, we don’t have medical knowledge that hints that the young people who get to critical condition have some a priori hidden condition that is expected to shorten their life anyhow. So we have to assume at this point that we would be losing many life-years under this strategy. See Figure 5 for distribution of mortality across age groups.
5. **Healthcare system strain will kill even more**. A herd immunity approach will strain the healthcare systems to its limits for many months. Healthcare personnel and resources will find themself recruited to care for the critical coronavirus patients, inevitably neglecting regular care. Many health situations that would normally end in recovery will end in otherwise avoidable death due to lack of care.
6. **Long term health damage**. It could well be that some portion of surviving patients would suffer long-term or permanent health consequences post recovery, with mid-term damage almost certain for those mechanically ventilated. Waiting for a vaccine could prevent it.

In light of the above, and given there appear to be other effective and relatively low-economical-damage containment strategies, it appears that sticking to containment strategies is more efficient at this stage.

## Discussion

Our results show that seemingly similar strategies in terms of the amount of population-business-days they allow, can have a very different epidemic supression outcome, based on the pattern the population is released. The more effective building blocks are a combination of both partial planned intermittent quaranties and partial population release, such as a two-group intermittent release or random shift. As such, these mixed intermittent approaches are better performing, and are reasonable to implement given they can be relaxed to allow some fixed population noncompliance - 10 percent in our example. Division to groups on a personal, rather than on a household basis make these building blocks easier to live with. It should be noted, however, that partial release schemes are sensitive to a special division into two groups that does not reduce interaction. For example, the resulting R of dividing the population into two groups on a city basis would resemble the resulting R of a single-group and less the R of two groups. In other words, if the division is not made in a way that shall reduce interaction we will get an R closer to that of a single group intermittent scheme.

Intermittent schemes without partial release of population seem less effective at reducing R compared to mixed schemes, though they are probably somewhat easier to enforce. Constant release of a fixed certain percent of population while keeping the rest quarantined also seems less effective too compared to mixed schemes where the population that is being released changes. Releasing the population, and then quarantining as the epidemic relapses seems less effective compared to planned intermittent release schemes that achieve better results while also less predictable business-wise.

The decision on using one building-block or the other is not just a numerical one, but should take into consideration other measures and factors including population compliance, effectiveness of economy, etc. The factors and their weight could change by population and region. Actual decisions are outside the scope of this paper.

A large untreated question is reopening schools. Unfortunately, there is just no data available ^35^ to make a truly informed decision, mostly because schools closed up very quickly globally with the appearance of the pandemic, with the exception of Sweden for the lower age-groups ^36^. Children are less symptomatic than adults ^37^. Furthermore, there is conflicting information on whether asymptomatic are less infectious ^38^ than symptomatic. Based on experience in China, Korea and Israel, it seems children contracted the virus less ^39^. According to Israeli ministry of health data ^40^, the percent of positive tests for children aged 0-9 years old is lower compared to adults and in the range of adults for ages 10-19, suggesting this phenomena of lower contraction in children is not merely an ascertainment bias due to milder symptoms. It could be argued the latter phenomena can be attributed to the fact the main importers of the virus were travelling adults and schools closed up relatively fast. In Sweden, the infection rates of children are also very low ^41^ - though testing was limited. In household secondary infection in China, children were less likely to get infected ^16,39^. In the municipality of Vo’, Italy, almost the entire municipality was tested twice as part of a survey in the early outbreak in Italy ^42^. While the infection prevalence was about 2.6%, none of the 234 children aged 0-10 were infected, despite some of them sharing a household with a case. According to Israeli ministry of health information, the education system accounted for only 3-11 percent of virus contraction. On top of things, and perhaps the bottom line - there is no real and reasonable solution for parents to go to work, if their children cannot go to their school - so opening of schools seems unavoidable. Limited mitigation can be partially achieved by noticing symptoms at school, and bi-daily measurements of body temperature for early detection of symptoms, as children tend to be less symptomatic.

We assume the epidemic shall last many months, and that schools will reopen before it’s eradicated. Therefore, we will have to accomodate for the school effect anyhow, whatever it is, so that R<1 despite schools being open. However, it could be that R would be very close to 1 after schools reopen, and so there could be benefits from reducing the number of new daily infections to a smaller level which would stay there after schools are reopened as R would be close to 1. Opening schools with our suggested strategy provides a layer of protection that would slow-down possible relapses.

## Summary

We compare key mitigation measures of SARS-CoV-2 spread, and compare various exit strategies building blocks. Our results stress the importance of not just the amount of population that is released, but also the pattern. Our findings demonstrate the importance of mixed strategies, as each strategy is effective through somewhat other means. Some of our mixed schemes can allow relatively convenient life while controlling for the epidemic spread.

Therefore, and given our results, we believe that pandemic can be controlled within a reasonable amount of time and at a reasonable socio-economic burden.

## Data Availability

No data is available

## Simulator and Underlying Assumptions

Our simulation is based on a SEIR (Susceptible, Exposed, Infectious, Recovered) agent-based model that we programmed in Python and adapted to Israeli population and parameters of SARS-CoV-2 epidemic. Therefore, in our database, we hold about 9 million rows, one for each resident of Israel, but for efficiency purposes we run the simulation on a subsampling of 1:10 on a reduced 900,000 representative lines. For each person, we hold an index, age, sex, and household identifier together with simulation data.

## Population Structure

We assumed each person is living with their children, unless they have children of their own. This construction resulted in a somewhat smaller household than real Israeli data. From Israeli bureau of statistics, the average Israeli household is 3.3 people ^43^, while in our simulation the average is 2.15, perhaps due to multi-generation households, or several families living together.

## Estimation of personal probability of death and mechanical ventilation

To mock coronavirus outcomes of mortality and mechanical ventilation for subjects, we used a combination of 3-year all-cause mortality predictor trained on Clalit healthcare data and have the results in cross-validation, and age-specific infection-fatality-rate (IFR) and ventilation estimates from the existing literature on SARS-CoV-2.

We manually fitted the mortality predictor to coronavirus mortality and mechanical ventilation probabilities as follows, where missing values were imputed by the reference value per appropriate age group.

| Age Group | Mean 3-Year Mortality Predictor value $P_i^M$ | SARS-CoV-2 Infection fatality rate <sup>34</sup> | Roughly Fitted Longevity Model $1-(1-P_i^M)^{0.25}$ [within <sup>34</sup> CI] |
| --- | --- | --- | --- |
| 0 to 9 | missing | 0.00161% | - |
| 10 to 19 | missing | 0.00695% | - |
| 20 to 29 | missing | 0.0309% | - |
| 30 to 39 | 0.2838% | 0.0844% | 0.107% |
| 40 to 49 | 0.6257% | 0.161% | 0.243% |
| 50 to 59 | 1.7059% | 0.595% | 0.678% |
| 60 to 69 | 3.6866% | 1.93% | 1.44% |
| 70 to 79 | 9.8188% | 4.28% | 4.00% |
| 80+ | 27.2394% | 7.8% | 11.99% |

| Age Group | Mean 3-Year Mortality Predictor value $P_i^M$ | SARS-CoV-2 Rate infected Hospitalized <sup>34</sup> | SARS-CoV-2 Rate Hospitalized Critical Care based on <sup>24</sup> | SARS-CoV-2 Rate infected Critical Care (mult. Prev. columns) | Roughly Fitted Longevity Model $1-(1-P_i^M)^{0.7}$<br>[within <sup>34</sup> CI mult. by rate] |
| --- | --- | --- | --- | --- | --- |
| 0 to 9 | missing | 0.1% | 5% | 0.005% |  |
| 10 to 19 | missing | 0.408% | 5% | 0.0204% |  |
| 20 to 29 | missing | 1.04% | 5% | 0.052% |  |
| 30 to 39 | 0.283% | 3.43% | 5% | 0.1715% | 0.187% |
| 40 to 49 | 0.625% | 4.25% | 6.3% | 0.2677% | 0.42% |
| 50 to 59 | 1.705% | 8.16% | 12.2% | 0.99% | 1.15% |
| 60 to 69 | 3.686% | 11.8% | 27.4% | 3.23% | 2.43% |
| 70 to 79 | 9.818% | 16.6% | 43.2% | 7.17% | 6.60% |
| 80+ | 27.239% | 18.4% | 70.9% | 13.04% | 19.05% |

## Temporal modeling

The simulation was based on the following temporal assumptions:

|  |  |
| --- | --- |
| Incubation: Time from infection to onset (of symptoms), given symptoms will occur. If symptoms do not occur we still simulate this time period for purposes of infectiousness which we assume is the same regardless. | gamma(5.807, scale=0.948), median 5.19, mean 5.5 days <sup>30</sup><br>We use gamma(5.807-3.27=2.537, scale=0.948) as time from infectiousness to symptoms onset. |
| Asymptomatic cases - will not develop symptoms | Assumed 0.2 <sup>44 18</sup><br>We set the time for being symptomatic as detailed below, but do not report the symptoms |
| Time from infection to being infectious | gamma(3.27, scale=0.948)<br>Based on <sup>15,23,38</sup>, the infectiousness starts at 2 to 2.3 days prior to symptoms, so we set the gamma parameters on the same scale as symptoms, but setting a=3.27, so mean is 3.09 - which is 2.41 days on average prior our chosen onset of symptoms. |
| Infectious period | gamma(9.21, scale=0.948), mean 8.737.<br>Accordingly, we use gamma(6.673, scale=0.948) post symptoms onset to end infectiousness, with a mean of 6.32 days post symptoms.<br>In <sup>23</sup>, 8 days are used post symptoms onset, and in <sup>15</sup> the effective range is 4-8 days post infectiousness onset - so our chosen value is in between. |
| Infectiousness intensity | Distributed as gamma(a=4.16, scale=0.948), where we scale the probability per person according to its total infectiousness period. So when calculating cdf(x), where d is the disease d starting with zero, and L is total infectiousness length. Then, $x=9.21(d/L)$.<br>Pre-symptomatic infection was reported as 12.6% <sup>45</sup>, 6.4% <sup>46</sup>, 44% <sup>23</sup> and 37% <sup>22</sup> of total pre- and post-symptomatic. We Average is 25%.<br>Post-symptomatic density should be about $(1-0.25)/0.25 = 3$ times the presymptomatic density.<br>A person turns symptomatic at 2.53 in our scale, the cdf at 2.53 is our chosen presymptomatic infection rate. The cdf for 2.53 is about 0.25. |
| Personal Probability of Mortality | $P^M=1-(1-P_i^M)^{0.25}$, see table above. |
| Personal Probability of ICU | $P^r = 1 - (1 - P_i^M)^{0.7}$, see table above. |
| Personal conditional Probability of ICU | 1 if Dies (Must spend the time in ICU)<br>If not dies, we draw a random variable with probability $(P^r - P^M)/(1 - P^M)$<br>(so total personal probability is $P^r$ under constraint of death only after ICU) |
| Time from onset to ICU | Gamma(10.5, scale=0.948), mean is 9.95, based on expert opinion slightly faster than <sup>47</sup>, For which median was 14.5 days<br>(If patient enters ICU, we draw a random variable under this probability + incubation time for the day they enter ICU from contraction) |
| Time in ICU | Assumed 10 days based on <sup>24,47</sup> |
| $R_{0\_Released}$ - the component of $R_0$ of non-household infections | 2.5<br>Results in $R_0$ of about 3.03 - which is a rough fit to Israeli epidemic early stage growth |
| $R_{0\_HH}$ - the component of $R_0$ attributed to household infections. | 0.485<br>Our estimate is based on <sup>13,16,39,48</sup>, which report $R_{0\_HH}$ of 0.4, 0.55, and 0.68, with average households of 4.28, 2.82, and 4.2 respectively. That is an attack rate of $0.4/3.28=0.121$, $0.55/1.82=0.3$, and $0.68/3.2=0.212$ respectively. The average being 0.211. As in Israel the average household is 3.3 persons, which minus the affected person is 2.3, we will use 0.485. |
| Percentage of symptomatic showing fever symptoms | 70% - Our estimate based on <sup>14</sup> reduced in light of Israeli Ministry of health epidemiology report. |
| Compliance Rate to self-household isolation | 95% upon appearance of fever, with a delay distribution of gamma(0.55, scale=0.948) - i.e., mean of half a day delay. Zero compliance otherwise. |
| Probability of those in quarantine to contract the virus from the outside world, regardless of reason (non compliance, human error, etc.) | 100 - fold reduction of the probability of released person |
| Infection from abroad | 10 randomly infected each day (if they have not contracted the virus already) for the first 30 days of simulation.<br>In reality this influx could continue assuming borders |
|  | remain open - and the economical cost of locking down borders or isolation incoming travellers is large. Still, we stopped after 30 days so we can better see the differences among various schemes. |

Note that available literature of these parameters has a wide confidence interval, which could affect our results.

## Simulation Temporal Course

The simulation is executed with a 1-day cycle simulating a year. Totals are counted for mortality, infections, ICU usage - all based on the above detailed assumptions. We did not take into account actual ICU resources in the simulation, but counted the needed resources and assumed they are available. It is up to the strategy to avoid a situation of exhausting resources.

At initialization, each subject receives concrete values of their personal disease course relative to infection, if they get infected. To that end we draw random variables of the relevant distribution detailed above. For each person we also hold a boolean flag indicating if they are infected, and a counter to count the relative number of days since each person’s infection. We also keep a flag to note if a person is in quarantine (isolation) or released. We assumed isolation is at home and with household members. People in ICU are considered removed and stop infecting others.

There are three types of infections modeled: infection of family members, infection of released population, and leakage infection to those in quarantine. The infection of each group is performed in complete mix, i.e., the probabilities to get infected by an infector are identical within a group, and the population within a group is uniformly infected based on that probability.

Daily infection of the released population is based on R_0_Released_ and the relative infectiousness for that day for an infector, whose product we denote as Φ_i_^d^. Thus, Φ_i_^d^ denotes the number of expected infectious interactions an infector *i* makes on day *d*, assuming no-one else is infected and all population is released. We define infectious interactions as the number of people that would be infected if no one else was infected yet. Then, when just a fraction *f* of the population is released, under complete mix assumption we assume the number of daily interactions per person drops by a factor *f*. Therefore, under complete mix, the number of infectious interactions an infector makes also drops by a factor *f*, and each infector shall now have only *f*·*Φ*_i_^d^ daily infectious interactions.

Interestingly, under the complete mix assumption, the probability of a released person to have an infectious interaction with a single infector is identical as in full release. However, there is a factor *f* less candidates to infect, so the total number of new cases drop by a factor of *f*. The probability of a released person to have an infectious interaction with an infected person is therefore equal to Φ_i_^d^/(T-1), where T is the total initial population - and one person is infecting the others. As T>>1 we neglect 1, and remain with infectious interaction probability of Φ_i_^d^/T.

Given there are C infectors released, under the complete mix assumption, the probability of a released person to be infected by one of them is:

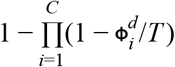

Using the approximation (1-1/x)^x^ is about e^-1^ for large x, where x=T/Φ_i_^d^, i.e., T>>Φ_i_^d^, it follows that the probability for a released individual to be infected in a day (if not already infected) is:

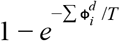

Our simulation tries to infect released individuals daily based on this probability. Note that when 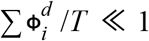, using the approximation that *e*^*x*^ = 1 + *x* for small x, the probability reduces to 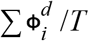. Though this approximation is used in classic analytical SIR/SEIR models, we did not have the need to use it in our simulations.

We can decompose R_0_ to contribution from household infections, and all the rest:R_0_=R_HH_+R_Rest_

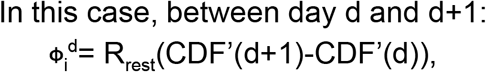

Where CDF’ is the cumulative density function of the chosen gamma probability for infectiousness scaled for the length of the infection period for this person. i.e., CDF’(d)=CDF(9.21(d/L)), where L is the personal length of infectiousness.

## Simulation Applied on a Strategy Building Block

The various strategies that are evaluated were fed separately into the simulator. A strategy can set for each person if they are quarantined or released, based on all prior information (age, sex, probabilities - but not actual random choices), and can measure actual ICU usage, and perform viral tests, require to put masks when out of home, etc.. The strategy is invoked each day and has a chance to intervene prior to the infections taking course for that day.

## Modelling Facial Masks

If two people are wearing masks, it reduces droplets and aerosol that contain the virus. Averaging the numbers in ^7^, we assume it reduces droplets and aerosol by about 29.1% to 18.2%, without change in the viral load, i.e., a mask on an affected individual reduces spread by a factor of 0.625 in a period of 30 minutes ^7^. Numbers are small so we assume about 30% spread both aerosols and droplets. We cannot make an accurate assumption on how much surgical masks shall reduce infection to a wearer of the mask, though based on ^49^ it would be about a six-fold protection factor. If we follow these numbers we expect infection to drop by a factor of 0.625/6 to 0.625, i.e., from 0.1 to 0.625. Another cause of inclarity is the possibility that droplets or aerosol are infecting through unprotected eyes, or other transmission from hands to the eyes ^50,51^, which are not protected by a surgical mask, and later accidental transfer from hands to mouth or eyes. The inclarity is further extended as good mask usage requires careful fitting of the masks, replacing them frequently enough, smart disposal together with additional protective measures ^52^.

It’s difficult to model long-exposure of several 30 minutes units, as we cannot tell if the percent of droplets are per-person due to mask-face fitting for example (although participants were corrected if they mis-wore the mask), or due to properties of the mask itself. If it’s mask properties then an hour worth exposure would still be infecting. e.g., even if we assume the extreme value for mask protection factor of 0.1, then for 5 hours the protection is only 1-(1-0.1)^2×5^=0.651, i.e., the protection is just 30% reduction in possibility of infection. So if surgical masks are useful, it’s probably protective for short and random exposure patterns. To provide a conservative yet useful estimate, we gace simulation results for protection levels of 10%, 30% and 50%.

## Estimation of impact on R_0_

SIR models typically state the S - Susceptible, E - exposed, I - Infecting, R - Removed, and N - total number of people. Then

dS/dt = -*β*SI/N^2^

dE/dt =*β*SI/N^2^-**κ**E

dI/dt = **κ**E - ***ν***I/N

dR/dt = ***ν***I/N

R_t_ is then (dE/dt + dI/dt + dR/dt)/(dR/dt), which is *β*S/***ν***N and R_0_ is then *β*/***ν***.

We can estimate R_0_ as if the S_0_ of the population is susceptible at start then: R_0_= ln(S_0_/S_∞_) / (1-S_∞_ / N), where we estimate S_∞_ by taking the uninfected number of uninfected people after a year, and S_0_ as the number of unaffected people at start of intervention. We do start intervening on day 14, but to allow washout of previous doing-nothing, we take S_0_ as day 35 (21 days washout). Taking S_0_ at day 35 also accommodates our simulation deliberate infection by 10 people a day during the first month.

## Notes

### Competing Interest Statement

The authors have declared no competing interest.

### Funding Statement

No external funding was received

